# Synergistic effects of natural products and commercial antibiotics—A mini–review 2010–2015

**DOI:** 10.1101/2020.09.01.20186353

**Authors:** Lucía Nitsch-Velásquez

## Abstract

1

**Context:** The ‘antimicrobial resistant era’ requires advances in the approaches and technologies to find new treatments. The enhancement of the antimicrobial activity of commercially available drugs (CADs) by natural products (NPs) has successful mixtures (*e.g*., clavulanic acid and amoxicillin).

**Objective:** To systematically review reports of synergistic effects of CADs and NPs against opportunistic microbial strains from 2010 to April 2016.

**Methods:** The databases and search engines PubMed, Medline, Scifinder, Scopus, ScienceDirect, Scholar Google were systematically searched. Among the keywords utilized were: synergistic effects natural products and antibioitcs, botanicals and antibiotics bioassays, plant extracts interaction with antibioitics and antibiotic adjuvant bioassays. Only synergistic results were tabulated and analyzed according to CADs, NPs and strains.

**Results:** A set of 76 studies that reported *in vitro* synergistic effects of CADs and NPs against gram–positive or gram–negative bacteria or fungi opportunistic strains was found. From the 60 reports on antibacterial adjuvants, the most frequent designs involved beta–lactamics or aminoglycosides against Methicillin Resistant *Staphylococcus aureus*. The assayed NPs encompassed extracts or fractions from 22 different species distributed worldwide (45% extracted with non–polar solvents) and 33 purified compounds (flavonoids, other polyphenols and alkaloids).

**Conclusions:** NPs as potential drug hits for antimicrobial adjuvants had been found and should continue in the drug discovery pipeline. The field certainly would benefit of advances in purification technologies, especially for polar extracts and bioassay platforms.

## Introduction

The challenge of the antimicrobial resistant era: For almost half a century the treatment scenario had been straightforward for a particular set of infectious diseases in humans, animals, and plants^1–3^.

The evolutionary pressure that represented the systematic administration of antimicrobials— together with its involuntary mismanagement due to a variety of reasons— in concert with the evolution intrinsic to microbe’s life^4^ has been yielding resistant strains^4,5^: microbes evolved diversifying their ‘hunt’ strategies^5^ (see examples in Fig. 1 .a and b.). The consolidation of the **antimicrobial resistance era** (ARE)^5–8^ represents one step backwards regarding the previous victory over the infectious diseases; this leads to the risk of the infectious diseases recurrence as the first cause of death worldwide. Beyond that, the ARE effects extrapolate to immuno– compromised patients^7,9^, veterinary and plant management. To tackle the ARE, the WHO had stablished guidelines and priorities, including the development of new drugs and pesticides^9^. So, it can be said that the combat has begun; as it is going on is becoming evident that the challenge is larger than we thought at the beginning, and more bioinspiration is welcomed^10^. It is on creative scaffolds where Nature plays on our side, since natural products often defy a medicinal chemist’s imagination.

**Figure 1:**
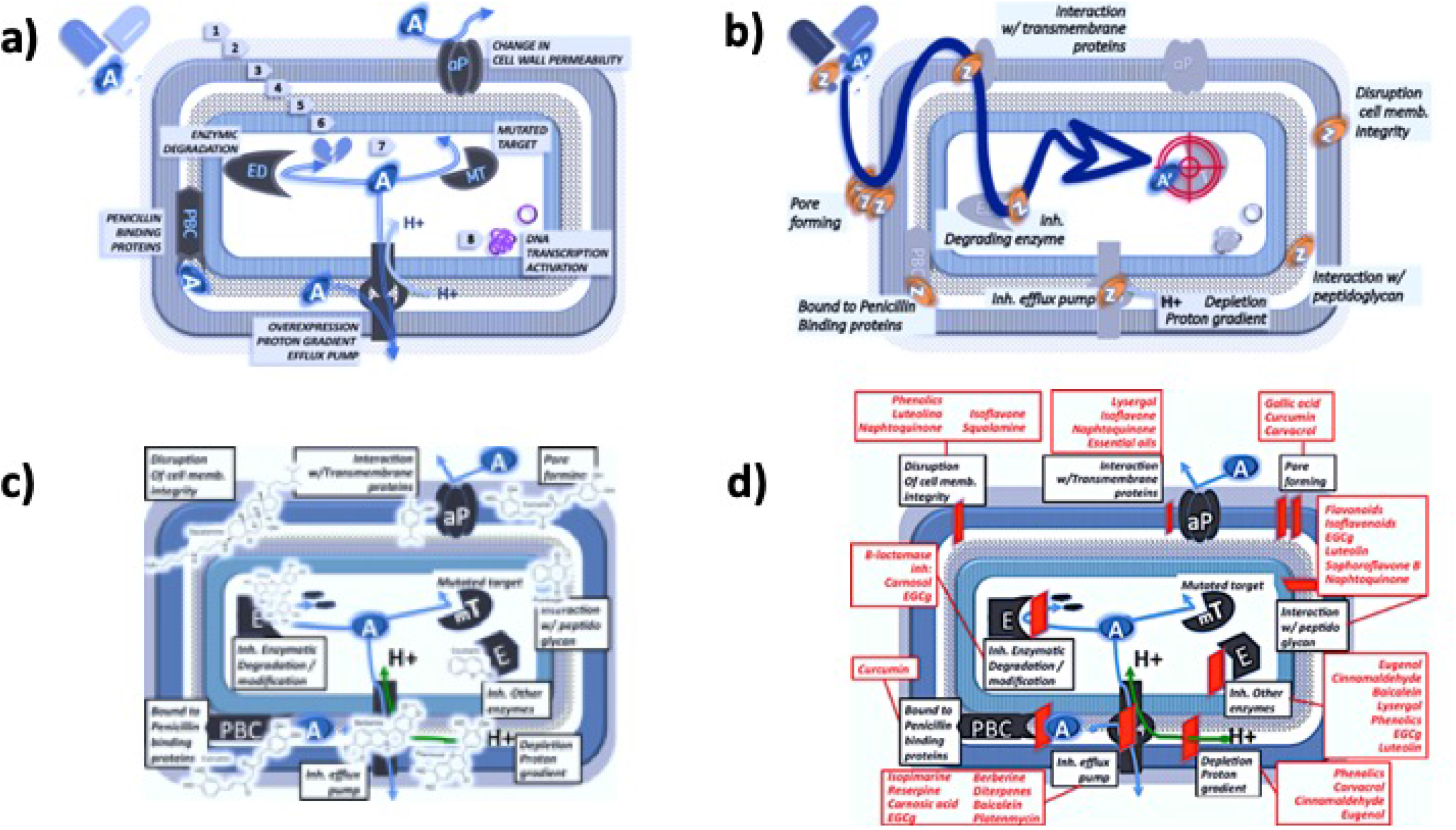
Depiction of examples of molecular mechanisms of antibiotic resistance and antibiotic adjuvant interactions with natural products at a cellular level. **a)** Examples of molecular mechanisms of antibiotic resistance, which are sites of potential interactions of phytochemicals with a generic resistant gram (-) bacteria^50,54,59,64–67,69–78^. . From extracellular space to inside: A = antibiotic, 1. extracellular space, 2. alginate, 3. outer cell membrane, 4. peri–plasmic space, 5. peptidoglycan wall, 6. inner cell membrane, 7. inner cellular space, 8. chromosomic and plasmid DNA. **b)** Examples of antimicrobial resistance mechanisms that can reduce the intrabacterial drug accumulation and adjuvant molecules with a generic gram–negative bacteria, such that facilitates the encounter of antibiotic with its target, which are sites of potential interactions with natural product compounds. In blue oval: A’ = a given antibiotic compound (no mutated target), In orange oval: Z = a given adjuvant agent(s), highlighted in red: the target of the antibiotics^50,50,54,54,59,59,64,64,65,65,66,66,67,67,69,69,70,70,71,71,72,72,73,73,74,74,75,75,76,76,77,77,78^. **c)** Potential interactions of NPs with subcellular components of a given Gram negative resistant bacteria^50,54,59,64–67,69–77^. **d)** Compilation of NPs and their proposed mechanism of action at subcellular level^50,54,59,64–67,69–77^.

The multi–targets therapies (MTT), as complex as they can be, are emerging as a viable option to be studied with rationale and random screenings (not necessarily with mechanisms of action based on one–drug–one–target or polypharmacology, *e.g*., ‘magic bullet drug’^11^). Combination therapy (CT), a type of MTT^11,12^ is one feasible option to combat the ARE (*e.g*., clavulanic acid–amoxicillin^13^ and treatments including immune system stimulaiton^14^. Phytopharmaceuticals **^A^** could be seen an MTT–CT option because a derivative from a given medicinal plant can contain a set of key components with a variety of targets, or one natural product can interact with several targets. But natural products as a CT option has a its own challenges to solve, *e.g*., expressly in quality control assurance, to innovate in the research pipelines and information organization to asure the achievement of drug candidates^16,17^.

A type of CT–research is the screening for natural products that can potentiate the antimicrobial activity of commercially available drugs (CADs), *e.g*., additive and synergistic effects. Natural products ranging from purified compounds to raw extracts, which can also have anti–inflammatory and immuno–stimulant activities. S cientific literature was reviewed to gain insight on the state of synergistic studies of NPs and commercially available drugs (CADs). Only reports that fell in the criteria below described were analyzed.

### 2.1 Literature Search Parameters

The literature search parameters were defined as following: **i) Databases and search engines:** National Center for Biotechnology Information (NCBI) – Pubmed Central,^18^ Scifinder,^19^ Scopus,^20^ Sciencedirect.^21^ In the cases in which the search engine also yielded recommended articles related to the found article, follow up of such studies was performed. **ii) Publication date:** in the range from 2010 up to April 2016. **iii) Targeted content:** Antimicrobial activity evaluation of commercially available drugs together with natural extracts against opportunistic microbes (CAD and NE–OM), such that yielded synergistic effects results which data analysis included either FICI or a statistical comparison between control and test groups. Also, analog results from testing the main component(s) of any given natural extract were also included. **iv) Keywords and phrases:** The keywords applied to start the literature search in the different databases and search engines were: synergistic effects natural products and antibiotics, botanicals and antibiotics bioassays, plant extracts interaction with antibiotics, and antibiotic adjuvant bioassays. **v) Exclusion criteria:** results classified as either antagonist, additive, or non–interaction effects of CADs and NPs tests; results classified as either enhancement or modulating effects of CADs and NPs tests, such that were reported without statistical analysis, such that it was not possible to conclude if synergistic effects were observed; only the NPs (either as extract or purified components from it) were tested for antimicrobial activity; mixture of two or more NPs, even if those yielded antimicrobial synergistic effects; mixture of two or more CADs, even if those yielded antimicrobial synergistic effects. **vi) Methodology:** Results obtained by the microdilution method were included; the design could be either by Fractional Inhibitory Combination Index—FICI determination, or if the differences between control and mixture were established as statistically significant **^B^**.

#### 2.1.1 Summary of the Literature Review Results

A total of 76 scientific reports of antimicrobial synergistic effects of NPs and CADs (NPs+CADs) were found, 89.5% are in PubMed database, being the years 2014 and 2015 with the highest publication frequency (see Fig. 2). Reports topics were distributed as follows: 60 related to antibacterials, four to antifungals and 12 to interaction mechanisms studies at cellular level. Studies were performed predominantly by interdepartmental collaborations distributed in four continents, only six reports involved international collaborations. The countries that mostly contributed were China, Korea and Brazil (range 10-15%). Asian countries were represented in about 56% of the articles. The articles were published in a variety of journals that belonged to different editorials.

**Figure 2:**
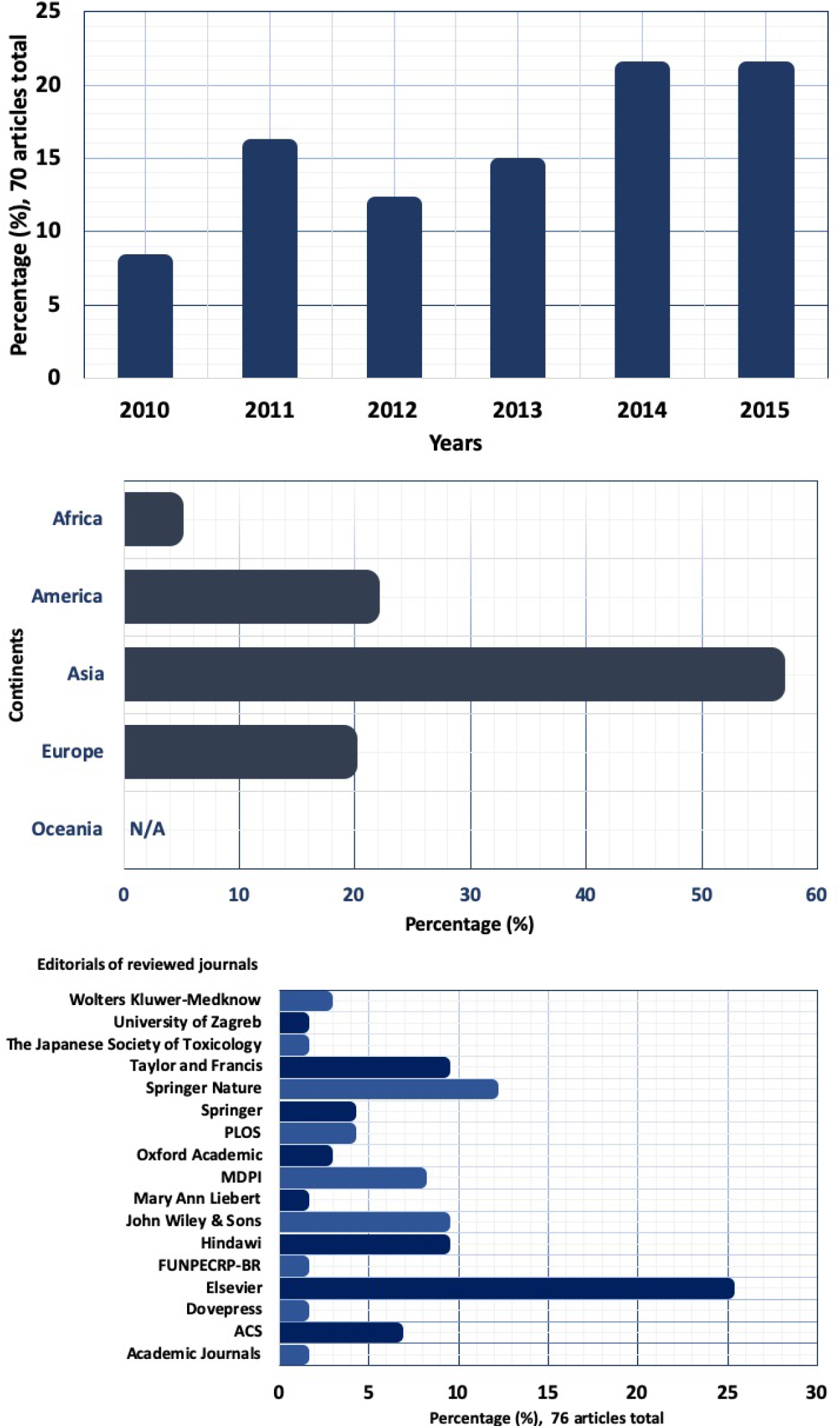
Distribution of the 76 reviewed publications by time, locations and editorials. **a)** Publication frequency during 2010 to 2016 (April). **b)** Approximated continental publication contribution. **c)** Article distribution in editorials (usually one editorial hosted more than one journal).

The 60 scientific reports of antibacterial adjuvants, published in 2010 to April 2016 are summarized in 3 and A and are compiled in Tables 1 and 2); in which the convergence of traditional medicinal knowledge and ‘Westernized medicine’ can be observed. For instance the evaluation of Sami–Hyanglyun–Hwan Korean formulation and ciprofloxacine^23^, or the study of traditional medicinal plants from Cameroon^24^. They are proof that the mining natural products *sensu lato* (NPs) **^C^**can still yield useful drug additives to treat resistant nosocomial bacteria (see Figs. 3 and A). Methicillin Resistant *Staphylococcus aureus*, MRSA, a strain for which new antibiotics are urgently needed^25^, was the most common target. The antimicrobial interactions of NPs+CADs were determined by the microdilution method, usually by the checkerboard method, applying serial dilutions in the twofold factor mode. (see Fig. A). The set of CADs encompassed medications that are under control for the emergence resistant strains against those antibiotics^26^; it also included antibiotics that the WHO consider as essential drugs in any health care system^27^. The bioassays either tested raw extracts or purified natural products. Among the natural sources investigated were plants across the globe, from which specific plant organs were extracted, usually selected based on ethnobotanical knowledge (see Figs. 4 and Table 1).

**Table 1:** Summary of plants screened for antibiotic modulatory effects of commercially available drugs, that yielded synergistic effects. For more details see text.^36,37^

| No | Plant extracted<br>(Plant family) | Potential<br>bioactive<br>compound<br>(Secondary<br>metabolite<br>type) | Plant<br>organ | Extraction<br>solvent | Microbial<br>strain | Antimicrobial<br>agents showing<br>synergistic<br>effects | Comments | Citation |
| --- | --- | --- | --- | --- | --- | --- | --- | --- |
| 1 | *Acacia mearnsii*<br>(*Mimosaceae*) | -- | B | P | SF (KZN)<br>PA (ATCC<br>19582)<br>EC (ATCC<br>25922)<br>BC (ATCC<br>10702)<br>SM (ATCC<br>9986)<br>EF (KZN)<br>SA (OKI)<br>ST (ATCC<br>13311)<br>ML<br>PV | ERY, CIP<br>ERY, TET,<br>CAM, KAN<br>TET, CIP, KAN<br>TET, CAM<br>TET, CIP<br>ERY, AMX,<br>CHL<br>ERY, TET,<br>AMX, CIP<br>TET, AMX, CIP,<br>NAL, KAN<br>AMX<br>ERY, TET,<br>AMX, CIP, KAN | antagonism<br>obsvd. | (Olajuyigbe &<br>Cooposamy, 2014) |
|  |  |  |  |  | KP (ATCC<br>10031)<br>PV (ATCC<br>6830)<br>SSn (ATCC<br>29930)<br>EC (ATCC<br>25922) | TET<br>TET, ERY<br>TET, ERY,<br>AMX, CAM<br>TET, CAM,<br>MTZ | antagonism<br>obsvd. | (Olajuyigbe &<br>Afolayan, 2012) |

**Cont. Table 1:** Summary of plants screened for antibiotic modulatory effects of commercially available drugs against nosocomial bacteria, that yielded synergistic effects.^24,38,79^

| No | Plant extracted<br>(Plant family) | Potential<br>bioactive<br>compound<br>(Secondary<br>metabolite<br>type) | Plant<br>organ | Extraction<br>solvent | Microbial<br>strain | Antimicrobial<br>agents showing<br>synergistic<br>effects | Comments | Citation |
| --- | --- | --- | --- | --- | --- | --- | --- | --- |
|  |  |  |  |  | EF (ATCC<br>29212)<br>EF (KZN)<br>BS (KZN) | MTZ, AMX<br>AMX<br>CAM |  |  |
| 2 | *Achyrocline*<br>*satureioides*<br>(*Asteraceae*) | 23-methyl-6-O-desmethyllauri-<br>cepyrone<br>(Xanthone) | L | P | SA | ERY, GEN | -- | (Joray et al., 2011) |
| 3 | *Aframomum*<br>*sulcatum*<br>(*Zingiberaceae*) | -- | S | W | SA<br>EC | CRO<br>NOR | antagonism<br>obsvd. other<br>strains (SS, SE, | (Njimoh et al., 2015) |
| 4 | *Albizia lebbek*<br>(*Fabaceae*) | -- | B | P | SA | AMP | antagonism<br>obsvd. other<br>strains (SS, EC,<br>KP) | (Njimoh et al., 2015) |
| 5 | *Alpinia officinarum*<br>(*Zingiberaceae*) | galangin<br>(Flavonoid) | R | N | SA (DMST<br>20651) | CEF | -- | (Eumkeb et al., 2010) |
| 6 | *Baillonella*<br>*toxisperma*<br>(*Sapotaceae*) | -- | B | P | SE | CRO, AMP | antagonism<br>obsvd. other<br>strains (EC, KP,<br>ST) | (Njimoh et al., 2015) |

**Cont. Table 1:**
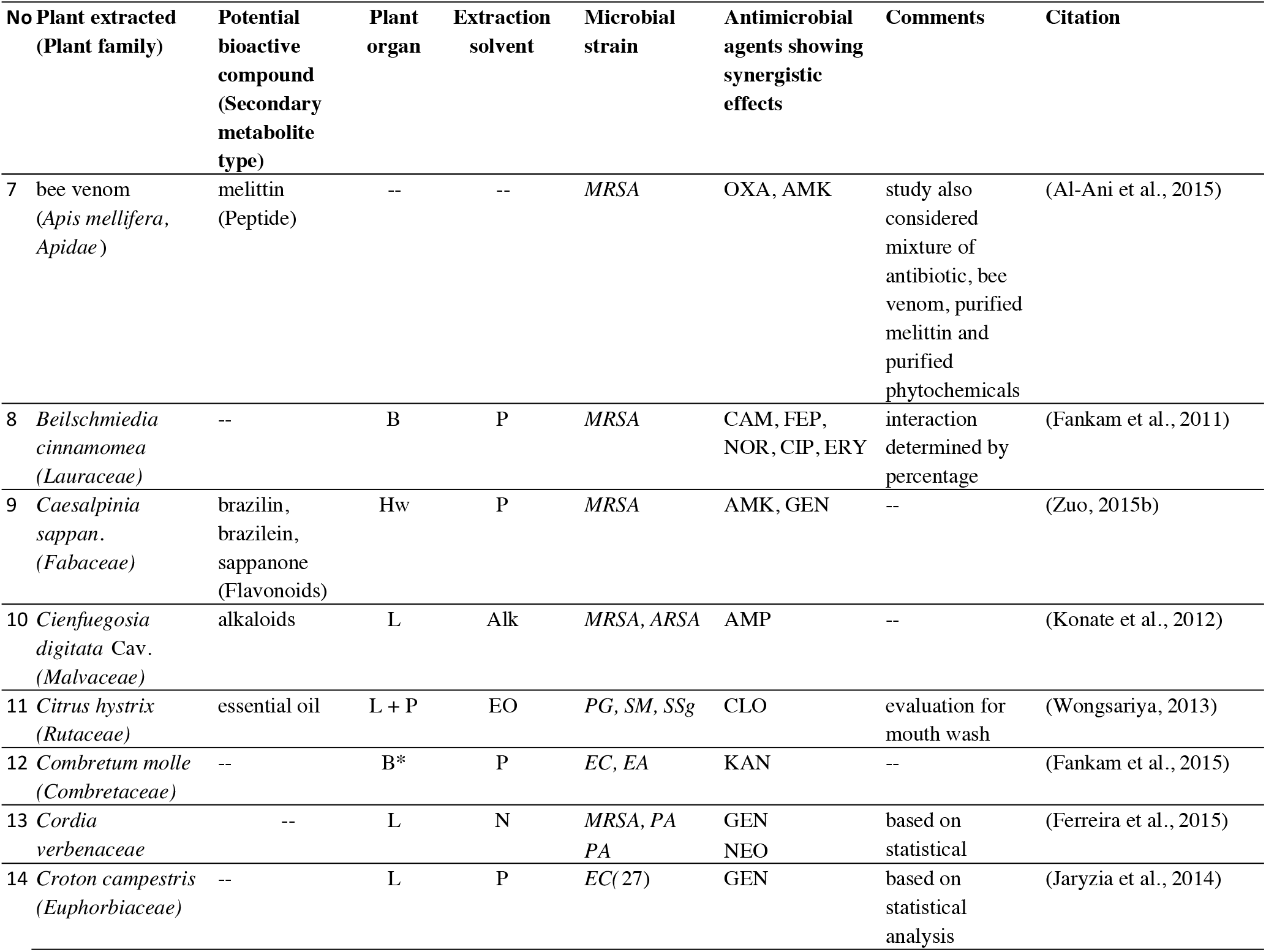
Summary of plants screened for antibiotic modulatory effects of commercially available drugs against nosocomial bacteria, that yielded synergistic effects^32,45,80–85^.

**Cont. Table 1:** Summary of plants screened for antibiotic modulatory effects of commercially available drugs against nosocomial bacteria, that yielded synergistic effects^24,39,45,86–91^.

| No | Plant extracted<br>(Plant family) | Potential<br>bioactive<br>compound<br>(Secondary<br>metabolite<br>type) | Plant<br>organ | Extraction<br>solvent | Microbial<br>strain | Antimicrobial<br>agents showing<br>synergistic<br>effects | Comments | Citation |
| --- | --- | --- | --- | --- | --- | --- | --- | --- |
| 15 | *Dichrostachys*<br>*glomerata*<br>(*Fabaceae*) | -- | Fr | P | *MRSA* | CAM, TET,<br>NOR | -- | (Fankam et al., 2011) |
| 16 | *Ecklonia cava*<br>(*Lessoniaceae*,<br>brown algae) | -- | "L" | NP, P | *MRSA* | AMP | -- | (Choi et al., 2010) |
| 17 | *Hibiscus sabdariffa*<br>(*Malvaceae*) | -- | Fl<br>(calyx<br>) | A | *HA* | CLA, MTZ | -- | (Hassan et al., 2016) |
| 18 | *Hypericum*<br>*japonicum*<br>(*Hypericaceae*) | taxifolin-7-O-alpha- L-<br>rhamnopyranosi<br>de (Flavonoid) | S | P | *MRSA* | CAZ, LVX | -- | (An et al., 2011) |
| 19 | *Hyptis martiusii*<br>(*Lamiaceae*) | -- | L | P | *MRSA* | KAN | -- | (Coutinho et al., 2010) |
| 20 | *Manuka honey* | -- | -- | -- | *MRSA* | TET, MUP | produced by bee<br>pollinization of<br>Manuka tree<br>flowers<br>(*Leptospermum*<br>*scoparium*) | (Jenkins & Cooper,<br>2012) |
| 21 | *Myracrodruon*<br>*urundeuva*<br>(*Anacardiaceae*) | -- | B | P | *SA, BS, KP,*<br>*EC* | GEN | -- | (Braz et al., 2015) |
|  |  |  | L | EO | *SA 358* | GEN | -- | (Figueredo et al., 2014) |
| 22 | *Nauclea pobeguinii*<br>(*Rubiaceae*) | -- | R | M | *SS*<br>*SE* | CRO, AMP,<br>AMX, NOR<br>AMP | -- | (Njimoh et al., 2015) |

**Cont. Table 1:**
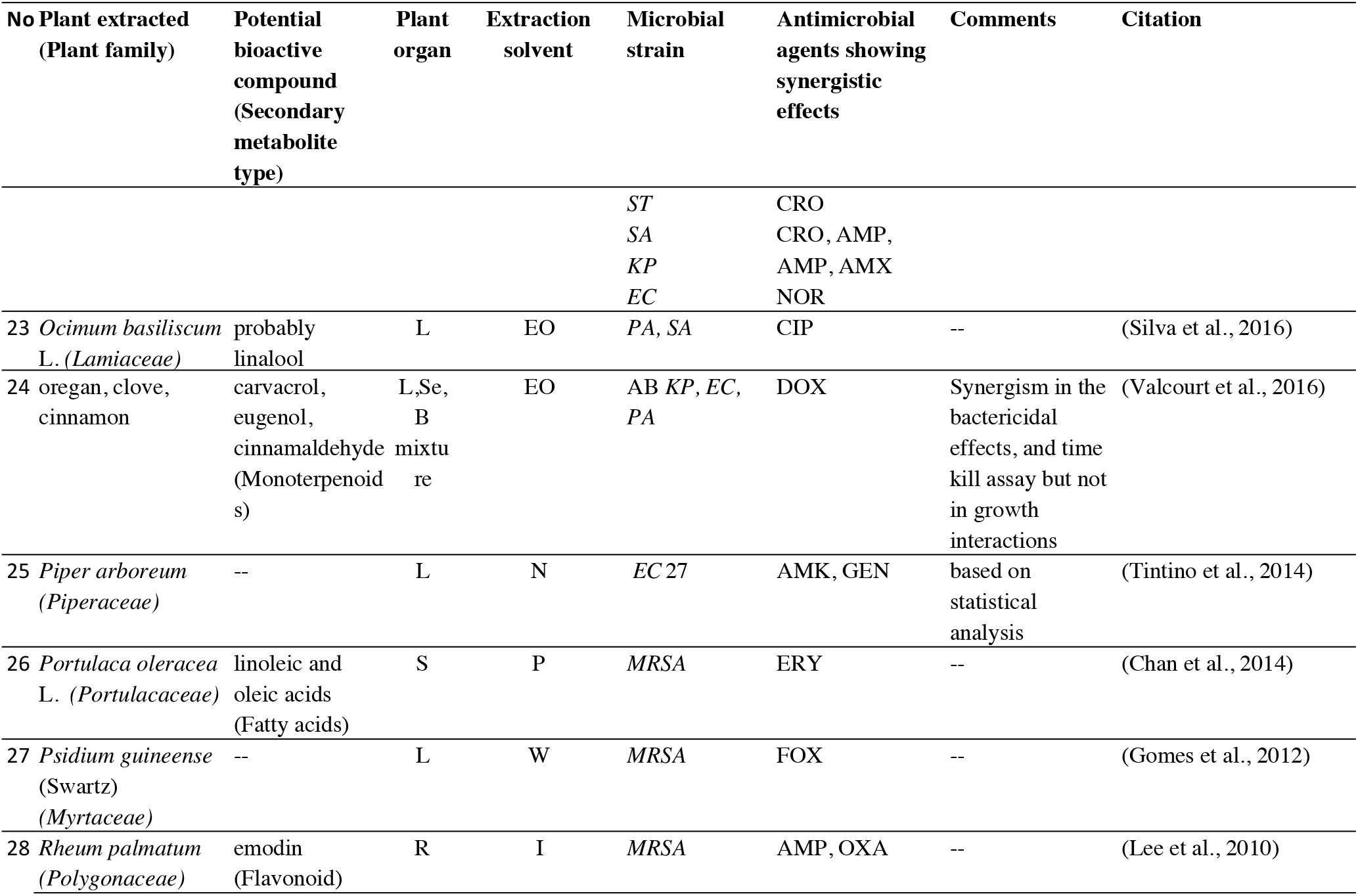
Summary of plants screened for antibiotic modulatory effects of commercially available drugs against nosocomial bacteria, that yielded synergistic effects^23,24,40,62,67,92–96^.

**Table 2:**
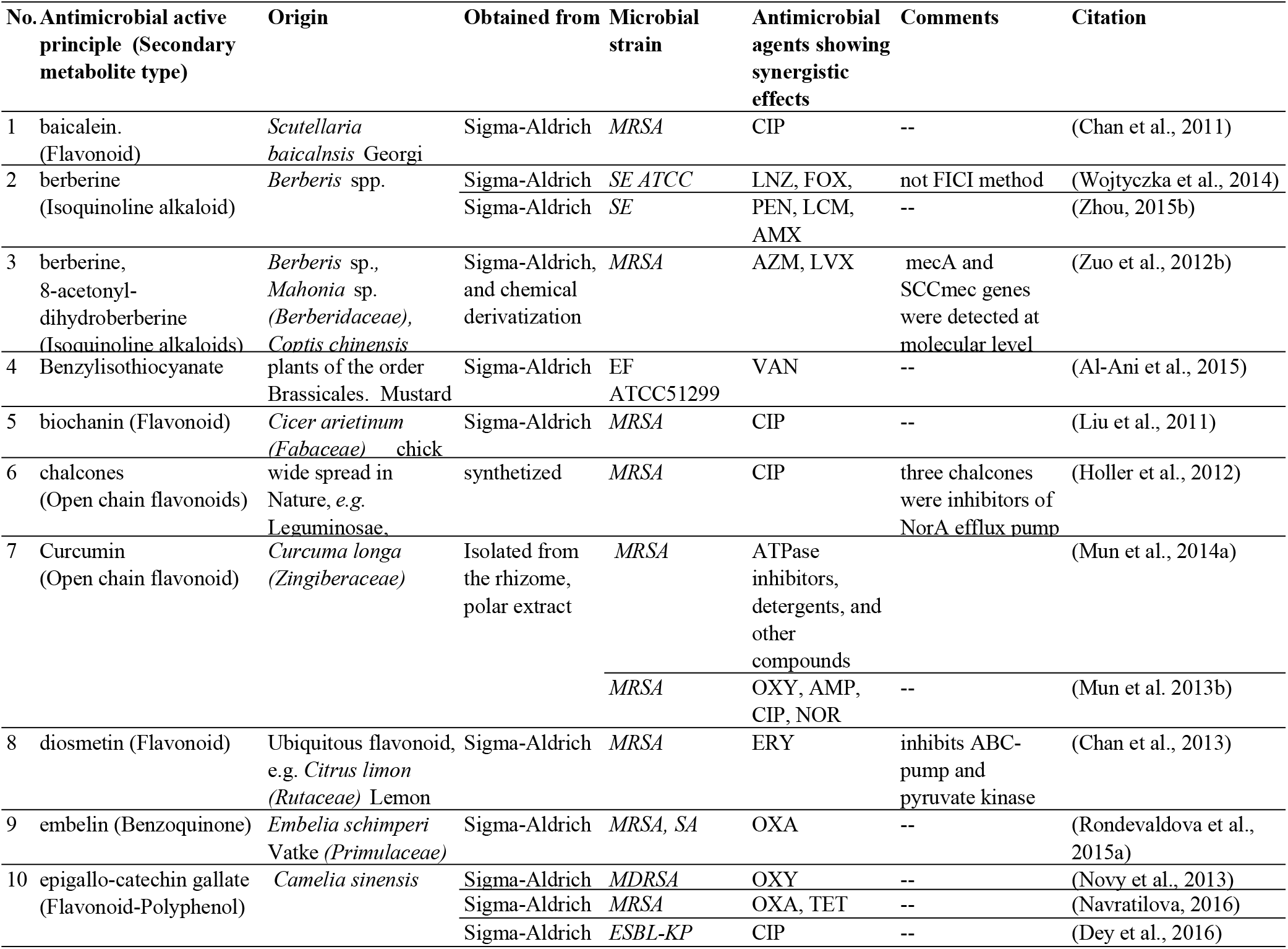
Summary of purified natural products screened for antibiotic modulatory effects of commercially available drugs against nosocomial bacteria, that yielded synergistic effects. For more details see text^32–34,46,47,51,70,71,97–101^.

| No. | Antimicrobial active principle (Secondary metabolite type) | Origin | Obtained from | Microbial strain | Antimicrobial agents showing synergistic effects | Comments | Citation |
| --- | --- | --- | --- | --- | --- | --- | --- |
| 1 | baicalein. (Flavonoid) | *Scutellaria baicalensis* Georgi | Sigma-Aldrich | *MRSA* | CIP | -- | (Chan et al., 2011) |
| 2 | berberine (Isoquinoline alkaloid) | *Berberis* spp. | Sigma-Aldrich | *SE ATCC* | LNZ, FOX, | not FICI method | (Wojtyczka et al., 2014) |
|  |  |  | Sigma-Aldrich | *SE* | PEN, LCM, AMX | -- | (Zhou, 2015b) |
| 3 | berberine, 8-acetyl-dihydroberberine (Isoquinoline alkaloids) | *Berberis* sp., *Mahonia* sp. (*Berberidaceae*), *Coptis chinensis* | Sigma-Aldrich, and chemical derivatization | *MRSA* | AZM, LVX | mecA and SCCmec genes were detected at molecular level | (Zuo et al., 2012b) |
| 4 | Benzylisothiocyanate | plants of the order Brassicales. Mustard | Sigma-Aldrich | EF ATCC51299 | VAN | -- | (Al-Ani et al., 2015) |
| 5 | biochanin (Flavonoid) | *Cicer arietinum* chick (*Fabaceae*) | Sigma-Aldrich | *MRSA* | CIP | -- | (Liu et al., 2011) |
| 6 | chalcones (Open chain flavonoids) | wide spread in Nature, e.g. Leguminosae. | synthesized | *MRSA* | CIP | three chalcones were inhibitors of NorA efflux pump | (Holler et al., 2012) |
| 7 | Curcumin (Open chain flavonoid) | *Curcuma longa* (*Zingiberaceae*) | Isolated from the rhizome, polar extract | *MRSA* | ATPase inhibitors, detergents, and other compounds |  | (Mun et al., 2014a) |
|  |  |  |  | *MRSA* | OXY, AMP, CIP, NOR | -- | (Mun et al. 2013b) |
| 8 | diosmetin (Flavonoid) | Ubiquitous flavonoid, e.g. *Citrus limon* (*Rutaceae*) Lemon | Sigma-Aldrich | *MRSA* | ERY | inhibits ABC-pump and pyruvate kinase | (Chan et al., 2013) |
| 9 | embelin (Benzoquinone) | *Embelia schimperi* Vatke (*Primulaceae*) | Sigma-Aldrich | *MRSA*, *SA* | OXA | -- | (Rondevaldova et al., 2015a) |
| 10 | epigallo-catechin gallate (Flavonoid-Polyphenol) | *Camelia sinensis* | Sigma-Aldrich | *MDRSA* | OXY | -- | (Novy et al., 2013) |
|  |  |  | Sigma-Aldrich | *MRSA* | OXA, TET | -- | (Navratilova, 2016) |
|  |  |  | Sigma-Aldrich | *ESBL-KP* | CIP | -- | (Dey et al., 2016) |

**Cont. Table 2:**
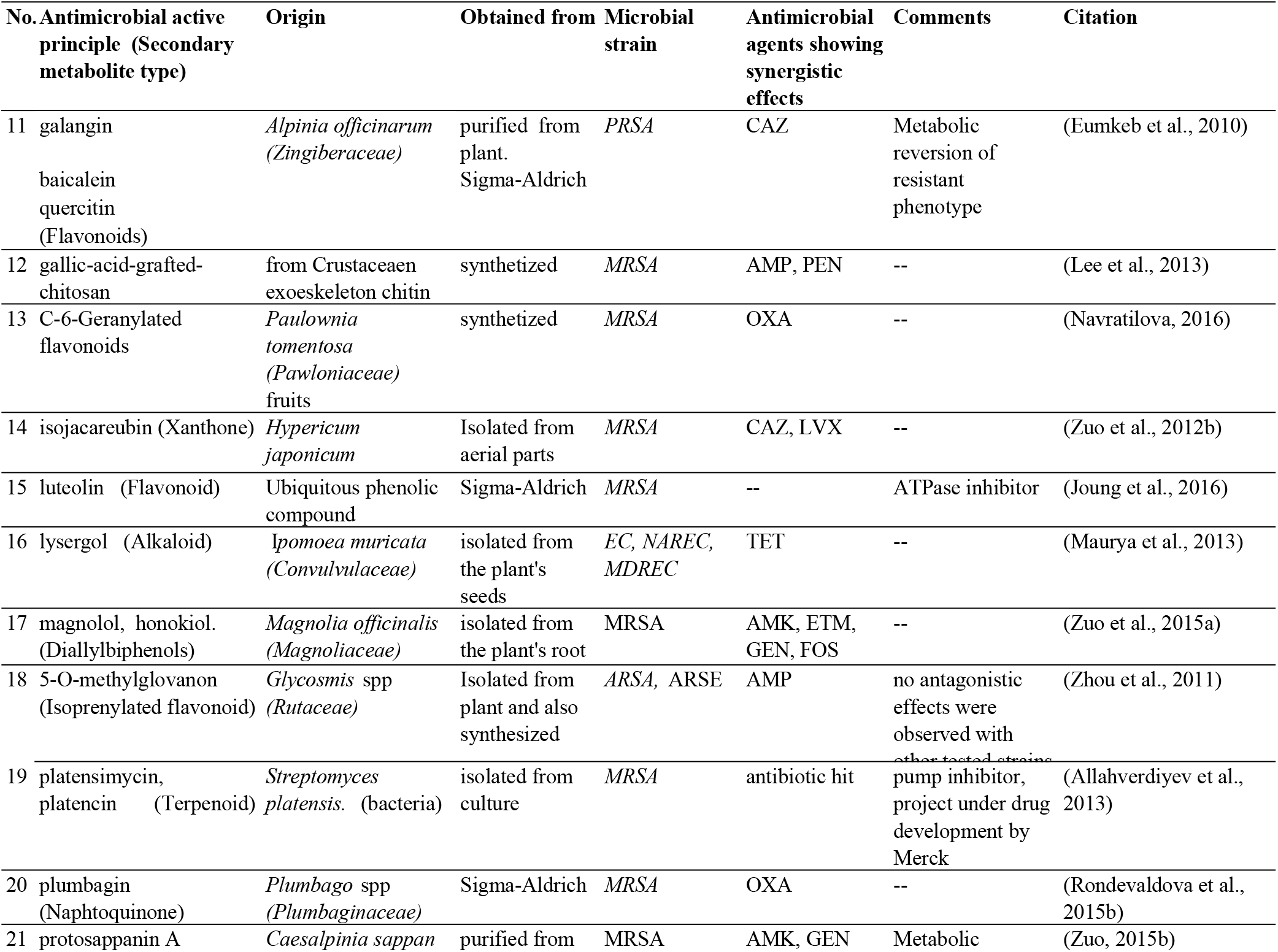
Summary of purified natural products screened for antibiotic modulatory effects of commercially available drugs against nosocomial bacteria, that yielded synergistic effects^38,47,48,60,64,76,80,99,101–103^.

**Cont. Table 2:** Summary of purified natural products screened for antibiotic modulatory effects of commercially available drugs against nosocomial bacteria, that yielded synergistic effects^35,55,63,69,75,80,104,104–110^.

| No. | Antimicrobial active principle (Secondary metabolite type) | Origin | Obtained from | Microbial strain | Antimicrobial agents showing synergistic effects | Comments | Citation |
| --- | --- | --- | --- | --- | --- | --- | --- |
|  | protosappanin B (Polyphenolic lactones) | (*Fabaceae*) | the heartwood |  |  | reversion of resistant |  |
| 24 | quercetin (Flavonoid) | Ubiquitous flavonoid | Sigma-Aldrich | *SA, MRSA* | FUS, MNO, RIF | -- | (Kyaw et al., 2012) |
| 23 | quercetin, morin, rutin (Flavonoids) | Ubiquitous phenolic compounds | Sigma-Aldrich | *MRSA, SA ATCC 43400* | AMX, AMP, CPD, CRO, IPM, MTH | -- | (Amin et al., 2015) |
| 22 | quercetin, luteolin, rhamnetin, octyl gallate (Flavonoids and other polyphenols) | Ubiquitous phenolic compounds | Sigma-Aldrich | *CP* | Ca 2+ modulators (isradipine, verapamil, thapsigargin) | -- | (Salin et al., 2011) |
| 25 | salvianolic acid (Trimer of a polyphenyl propanoic acids) | *Salvia miltiorrhiza* Bunge (*Lamiaceae*) | Shanghai Greenvalley Pharmaceutical Co., Ltd, Shanghai, Chin | *MRSA* | AMP, AMK | -- | (Liu et al., 2015) |
| 26 | sanguinarine (Benzophenanthridine alkaloid) | *Sanguinaria canadensis* (*Papaveraceae*), root | Sigma-Aldrich | *MRSA* | CIP, NOR, OXA | -- | (Obiang-Obounou et al., 2011) |
| 27 | Silibinin (Flavonolignan) | *Silybum marianum* (*Asteraceae*), seeds | Sigma-Aldrich | *MRSA* | OXA, AMP | -- | (Kang et al., 2001) |
| 28 | sodium new houttuynfonate (Sulfonic acid, surfactant) | *Houttuynia cordata* (*Saururaceae*) | isolated from plant | *MRSA* | OXA, CF, MEM, NTM | -- | (Lu et al., 2013) |
| 29 | sophoraflavanone B (Flavonoid) | *Desmodium caudatum* (*Fabaceae*) | Sigma-Aldrich | *MRSA* | AMP, OXA, GEN, CIP, NOR | -- | (Mun et al., 2013a) |

**Figure 3:**
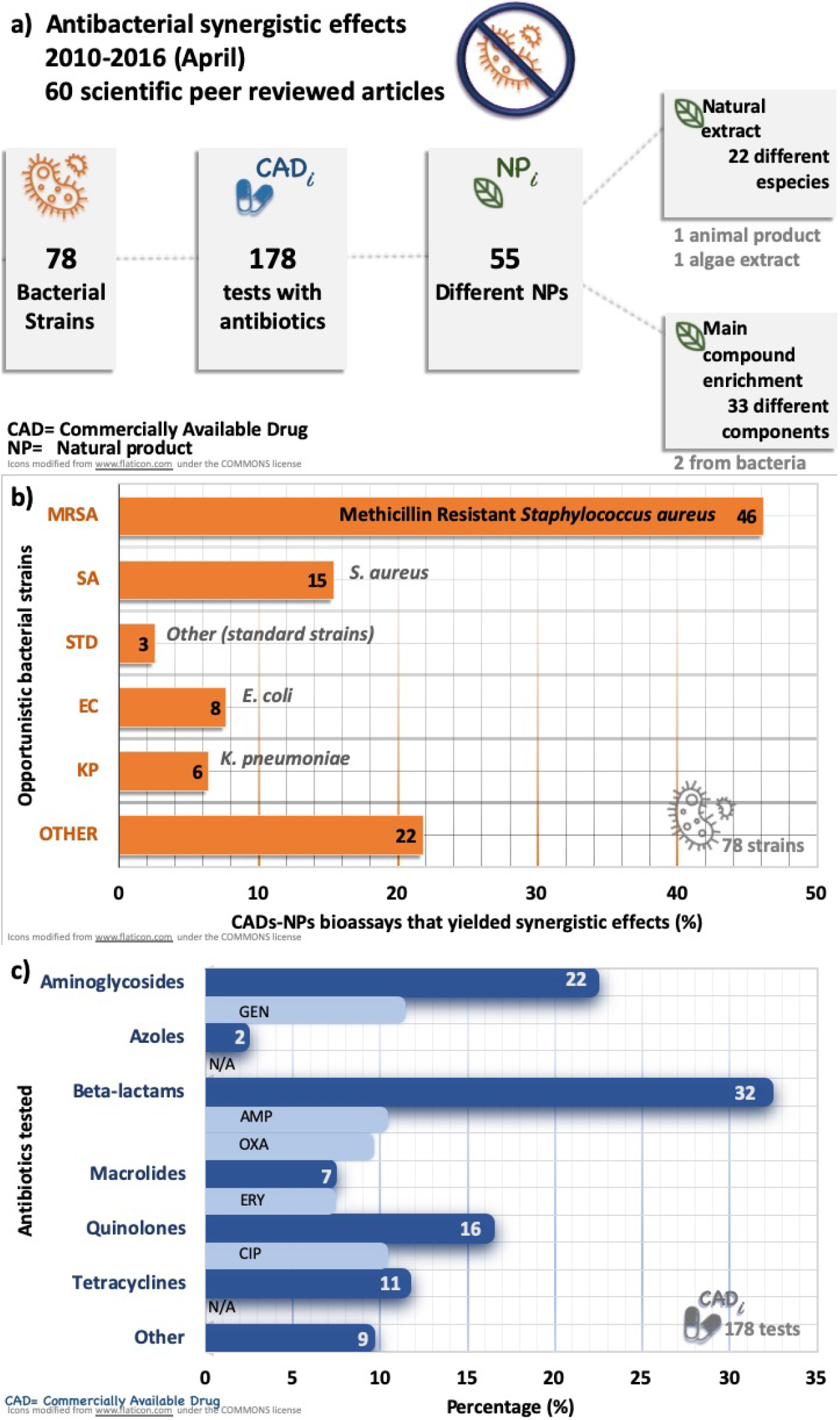
Summary of results of 64 studies of antibiotic synergistic assays of NPs and CADs **a)** Over all results were categorized by antibiotics, strains, and NPs. **b)** Distribution of bacterial strains that were susceptible to the mixture of NPs and CADs at a synergistic level. **c)** Distribution of antibiotics families that yielded synergistic effects with different NPs.

Tables 1 and 2 compile the organism’s family and scientific names, potential bioactive compound, organ extracted, polarity of the extracting solvent, the bacterial strains and antibiotics that yielded synergistic effects, comments, and the respective citation. General comments of three parameters are given below: commercially available drugs tested, bacterial strains assayed, and natural products tested. **Commercially available drugs (CADs):** from the 47 different drugs tested; 46 were antibiotics from different antibiotic families, and one drug was an antimicrobial with dual activity against bacteria and protozoa (see Fig. 3. In all the studies, the antimicrobial agents were purchased as at least 95% pure, and not as pharmaceutical formulation. The most frequently assayed antibiotics were (the antibiotic’s family is given in parenthesis): ampicillin (beta–lactams), ciprofloxacin (quinolones), and gentamicin (aminoglycosides). They are in the list for essential drugs in a health care system, recommended by the WHO and under the resistant surveillance projects by CDC and the British Society for Antimicrobial Chemotherapy, BSAC as well.

The criteria for antibiotic selection varied from study to study, and often was not clearly stated In general, the most common motivations described in the articles’ introductions were: **a)** the antibiotics’ toxicity would be reduced by lowering effective concentration **^D^**, **b)** the economical burden associated to infectious diseases would be alleviated by shortening the dosages and hospitalization expenses, and **c)** the reverse the antibacterial resistance can potentially be achieved by exploring natural products bioactivity.

**Bacterial strains** usually were clinical isolates with a specific resistance (no full antibiogram reported). In the reviewed reports, the utilization of standard strains were the exception more than the rule (3% of the tested strains)^32–40^. In almost all the cases, resistant opportunistic bacteria were the target, the most frequently strain tested were clinical isolates of MRSA. MRSA is in the WHO list of High Priority Bacteria that requires research and development of new antibiotics^25^, and the CDC classifies MRSA as Serious Threat Level^9^. In 2014, MRSA was included among the **US government National Targets for Combating Antibiotic-Resistant Bacteria**, aiming to reduce by half (to—at least—50%) the bloodstream infections caused by MRSA^41^. *Shigella flexeneri*, an intestinal pathogen, was the only pathogenic bacteria tested in the set of publications reviewed.

Several of the studied strains belong to the so–called ESKAPE pathogens species (**E***nterococcus faecium*, **S***taphylococcus aureus*, **K***lebsiella pneumoniae*, **A***cinetobacter baumanii*, **P***seudomonas aeruginosa*, **E***nterobacter*)^42^. The ESKAPE strains are a set of antibiotic–resistant pathogenic bacteria that represents new paradigms regarding pathogenesis, transmission, and resistance^42^ Opportunistic bacteria not only represents a challenge in the ARE but can serve as model for other pathogenic strains that affect not only humans but animals and plants. Further tests of the extracts that yielded synergistic effects with CADs against ESKAPE pathogens would allow to asses their potential as drug candidates, as well as cytotoxicity assays. Gram–negative bacteria still represents a medicinal chemistry challenge. Even though gram–negative strains were included in the assays, synergistic effects were found mostly for gram–positive bacteria. The alkaloids lysergol and squalamina, flavonoids quercetin, luetolin, rhamnetin, and other polyphenols octyl gallate, benzylisothiocyanate, epigallo–catechin gallate, and the vitamin alpha–tocopherol yielded synergistic effects against gram–negative bacteria.

At the same time, the cases of selective synergistic and antagonistic effects^36,37^ can be of interest for biofermentation purposes. The characterization of the tested strains should be pursued to be as complete as possible. Most of the studies were exploratory and further microbiology tests could be performed in collaborations with international collaborations (for instance CO–ADD^43^ initiative could be an interesting option).

**Natural products (NPs)** were classified in two categories: raw extracts and main component of a given botanical extract. which were (see Fig. A): *i)* purified from the raw extract and whose molecular structure was elucidated, *ii)* prepared by total synthesis, or *iii)* commercially available NPs (see Table 2). NPs derived from other kingdoms were less frequently found, *e.g*., honey, algae, bacterial derived drug leads: platencimicyn and platencin.

**Raw extracts:** The compiled descriptors were: organism’s family and scientific names, organ extracted, and polarity of the extracting solvent (see Fig. 4). The **plant organ** most frequently analyzed were the stems, the aerial parts extracted summed up to 35% (see Fig. 4) Reports exploring flower extracts were not found; while the phenological state (*e.g*., seeding, flowering) or collection date were uncommonly reported.

**Figure 4:**
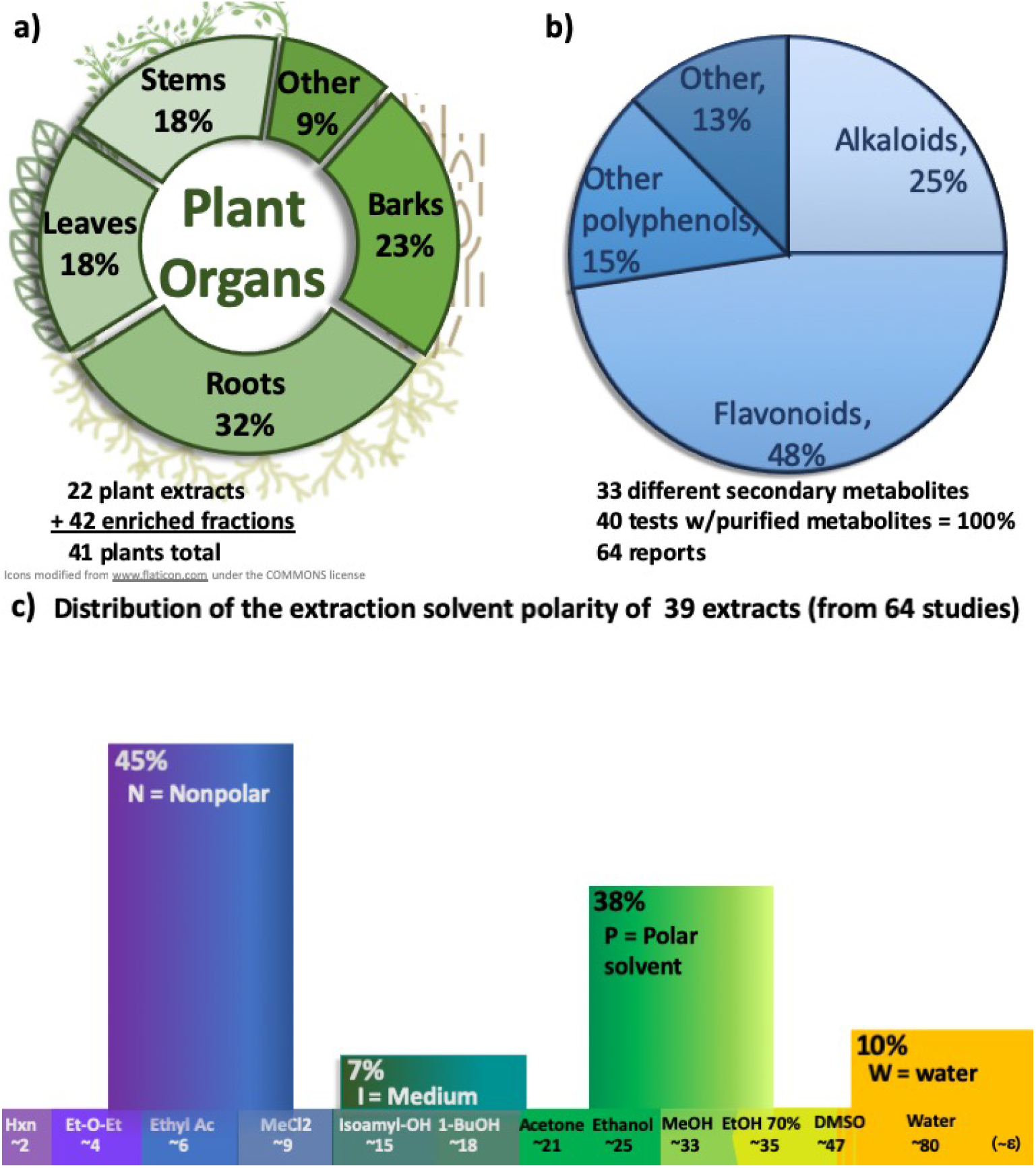
**a)** Plant organs extracted. **b)** Distribution of purified natural products assayed in the set of reviewed scientific reports (that yielded synergistic effects with antibiotics). **c)** Extraction solvents, ordered according to their dielectric constant (*ε*).

The *natural source* and *extract’s polarity* were usually chosen based on references from traditional medicinal knowledge or ethnopharmacology. Other criteria were the follow up of a given specie, genera, or family; and one study aimed the exploration of side products from industrial processes of an essential oil producing plant^24^.

The **extraction procedure** was described in all the studies (maceration, infusion, reflux, or soxhlet methods). The majority of tested extracts were fractions (or subfractions) extracted with low–polarity solvents (see Fig. A.d). For instance, the alcoholic extracs were further fractionated, the non–polar subfraction was then bioassayed (*e.g*., ethyl acetate sub–fraction).**^E^**

Several techniques were utilized to analyze the extracts’ composition, *e.g*., a thin layer chromatography, high performance liquid chromatography coupled to mass spectrometry. However, such analyses were not consistently included.

Polar extracts, the less utilized in the given reports set, can contain bioactive compounds that are already soluble^24,36,37,45^. However, the intricate nature, especially of aqueous extracts, has been limiting their exploitation. It is important to develop technologies that can allow for a more feasible purification process of polar extracts.

Chemical analysis and further bioactivity characterization should be pursued for the extracts in Tables ?? to 2. For example, the activity of *Acacia mearnsii* bark polar–extract had been explored against a variety of ESKAPE strains^36,37^, further characterization may lead to find drug hits and understanding of the antagonistic effects observed for some of the strains studied.

**Main components of a given botanical extract or purified NPs** whose molecular structure have been elucidated were described including *i* common chemical name, *ii* compound classification according to the secondary metabolites family, *iii* plant of origin, *iv* if the substance was purified from the raw extract, prepared, or commercially available (see Tables 1 and 2). The family most frequently tested were flavonoids (mostly chalcones and flavanones). To date some of them are included in dietary supplements that are not necessarily approved by the Food and Drug Administration, FDA. For instance, quercetin in vitamin C supplements that claim to boost the immune response^14^. Flavonoids show a variety of bioactivities —besides antibacterial— with a mid–intensity effect, *i.e*. anti–inflammatory and antiviral activities. Efforts toward understand their mechanism of action at a molecular level are ongoing^12^.

The adjuvant antimicrobial activity space for *alkaloidal family* was less explored. The 25% of the NPs families explored for that bioactivity (see Fig. A). Three alkaloids were clearly defined as druggable structures: berberine, platensymicin, platencin (see Tables 1 and 2) . These three alkaloids showed stronger adjuvant potency with a more specific effect than the set of flavonoids tested^33,46–48^, the latter two are being considered to be antibiotics *per se* and are under further development by Merck pharmaceuticals^48^ (see Table 2). The mechanism of action of these three compounds has been proposed to be orthogonal to those of the CADs. Further functionalization of their alkaloidal skeletons may allow improvement of their medicinal chemistry^33,46–48^.

The dominant *supplier of natural products*, by either natural extract purification or organic synthesis preparation, was Sigma–Aldrich company.

It is worth to mention that the aqueous extract obtained as a side product of the industrial essential oil extraction, the so–called the hydrolate, was tested^24^, which fits in the bioprospecting framework.

Successful examples of CADs+NPs, at in vitro level, hint that the enhancement of the antibacterial activity of CADs is a worthy approach to keep exploring and expanding. As it is further exploited, some challenges need to be addressed (or taken in account): **a)** A given extract can yield synergistic effects for some strains while antagonistic for other strains. **b)** The reproducibility is an issue related to the intrinsic complexity of natural extracts. **c)** The information included in the scientific reports can be more consistent across literature, *e.g*., extract’s composition, control experiments, strain profile. **d)** The need of widespread a high throughput analysis platform to speed up the analysis in each stage of the NPDD. **e)** Funding related to bioprospecting studies, as well as human resources, are required. **f)** Innovative approaches, aiming to increase the success rate of plausible medications derived from NPs and CADs mixtures need to keep developing.

#### 2.1.2 Examples of NPs Mechanisms of Action That are Orthogonal to the Antibiotics’ Mechanism of Action

Proposed interactions of NPs with bacterial components are depicted in Figs. 1 and 1. Curcumin, epigallocatechin gallate (EGCg) and berberine are examples of natural products whose mechanism of action have been frequently studied at sub–cellular level. The main component in the curcuma spice, curcumin, disrupted the bacterial cell membrane, probably by intercalating in the lipid bilayer^49–51^. The main antioxidant component in green tea, EGCg, inhibited beta–lactamase (an antibiotic degrading enzyme); EGCg interacts with cell membrane components, disrupting the cell membrane integrity^50,52–60^. The alkaloid berberin inhibited the efflux pumps with medium specificity—a resistance mechanism— enabling the antibiotics to remain longer periods inside the bacteria to interact with the desired target^46,56,61–63^. Bacterial cell membrane disruption is a non–specific interaction, saponins, other polyphenols and flavonoids^64^, and essential oils can exert this effect as well^36,65^.

Bacteriostatic effects of polyphenolic compounds such as coumarins, gallic acid, and flavonoids are attributed to: *i* to precipitate proteins, a non specific interaction that reduces the bacterial metabolism^65,66^; *ii* to act as enzymatic reversible inhibitors^65,66^, reducing the bacterial metabolism, especially if the targets are housekeeping processes; or *iii* to deplete the bacterial proton motive force and electron flow, thus reducing the amount of the biochemical energy available, which can induce a non–reproductive state in the bacterial metabolism^67^. As more information on these mechanisms of actions is gathered, the medicinal chemistry of NPs as drug candidates can be improved. Assays aiming a high throughput approach for the exploration of NPs mechanisms of action against bacteria have been developed, for instance the NPs interactions with peptidoglycan, ATPases inhibitors, efflux pump inhibitors, membrane disruption^50,54^. Studies on inhibitors of bacterial biofilm formation^68^ requires more attention—from the NPs perspective.

**Antifungal synergistic effects:** four reports were found testing a total of 12 opportunistic strains including filamentous fungi (see Table 3). Three extracts and two purified natural products yielded synergistic effects with azole type antifungals.

**Table 3:**
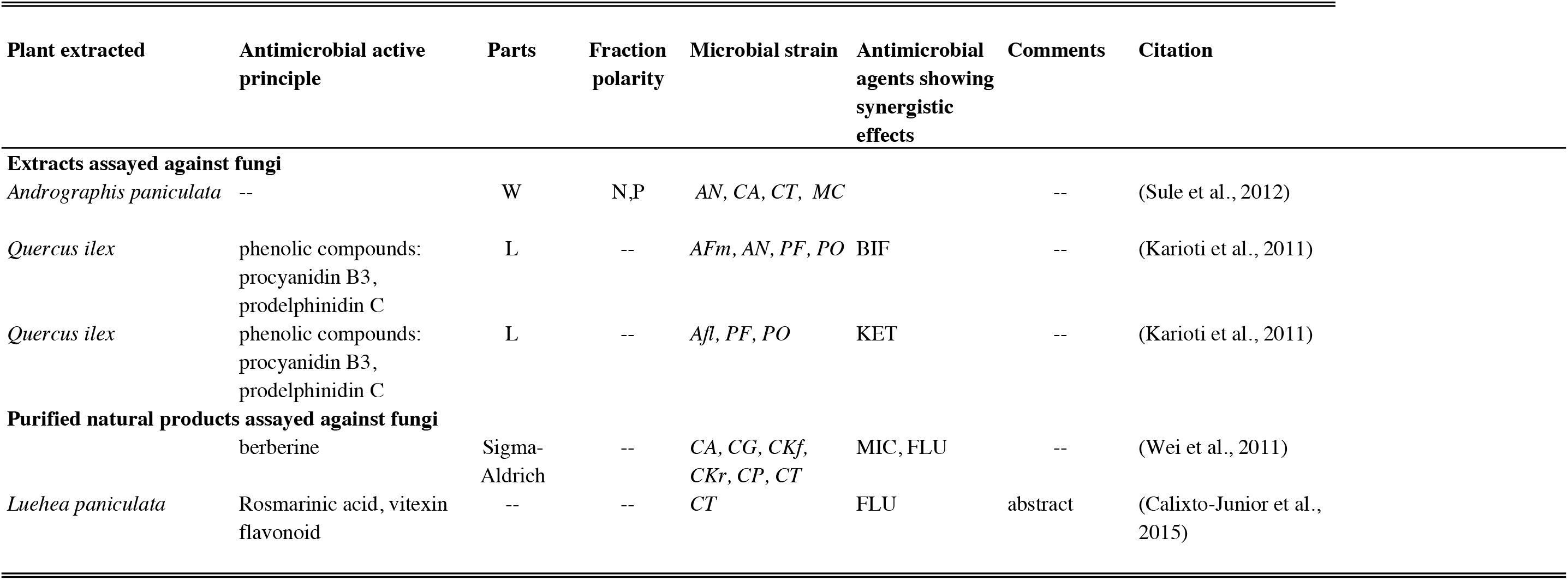
Summary of plants screened for antimicrobial modulatory effects of commercially available drugs, that yielded synergistic effects against fungi^111–114^. Strains: AFl=*Aspergillus flavus*, AFm=*Aspergillus fumigatus*, AN=*Aspergillus niger* CA=*Candida albicans*, CG=*Candida glabrata*, CKr=*Candida krusei*, CKf=*Candida kefyr*, CP=*Candida parapsilosis*, CT=*Candida t*, MC=*Microsporus canis*, PN=*Penicillum funiculosum*, PO=*Penicillium ochrochloron* Antimicrobials: BIF=bifonazole, FLU=fluconazole, KET=ketoconazole, MIC=miconazole.

| Plant extracted | Antimicrobial active principle | Parts | Fraction polarity | Microbial strain | Antimicrobial agents showing synergistic effects | Comments | Citation |
| --- | --- | --- | --- | --- | --- | --- | --- |
| **Extracts assayed against fungi** |  |  |  |  |  |  |  |
| *Andrographis paniculata* | -- | W | N,P | AN, CA, CT, MC |  | -- | (Sule et al., 2012) |
| *Quercus ilex* | phenolic compounds: procyanidin B3, prodelphinidin C | L | -- | AFm, AN, PF, PO | BIF | -- | (Karioti et al., 2011) |
| *Quercus ilex* | phenolic compounds: procyanidin B3, prodelphinidin C | L | -- | Afl, PF, PO | KET | -- | (Karioti et al., 2011) |
| **Purified natural products assayed against fungi** |  |  |  |  |  |  |  |
|  | berberine | Sigma-Aldrich | -- | CA, CG, CKf, CKr, CP, CT | MIC, FLU | -- | (Wei et al., 2011) |
| *Luehea paniculata* | Rosmarinic acid, vitexin flavonoid | -- | -- | CT | FLU | abstract | (Calixto-Junior et al., 2015) |

## 3 Final remarks and comments

The field of antimicrobial synergistic effects is active and yielding drug hits and extracts with promising bioactivities and mechanisms of action. In order to advance in the drug discovery research pipeline is important to pursue the full characterization of the natural products and strains tested; for which inter– and multi–disciplinary collaborations at local and international level are needed and funding agencies institutions should enhance support, not only by the grants *per se* but by facilitating the interactions of scientist from different areas such that more collaborations can emerge. Interactions of this scientific community with industries that utilize plants as raw materials can help to identify potential niches for further plant waste exploitation, contributing to form circular economies in the agroindustry.

## Data Availability

Compiled information from studies is included in tables.

## Acknowledgments

Thanks to Dr. Troy Wood, Dr. Anthony Campagnari and Dr. Nicole Luke (UB), Dr. Brooke (UVG) for sharing their expertise.

## A Compilation of the Reports of Synergistic Effects of NPs and CADs

The compiled reports of binary mixtures of natural products and commercially available drugs, NPs+CADs, that yielded synergistic effects against opportunistic bacteria were divided in two types of natural products, plant extracts (Table 1) and purified natural products (Table 2). A section of fungicide activity is in Table 3.

## Footnotes

A The Indian governmental drug regulation agency defines phytopharmaceuticals as a fraction of an extract from a given medicinal plant or its parts. Such that the fraction has been purified and standardized in its composition of at least four compounds, which have been detected and quantified. It encompasses the preparation administration for internal or external use in humans or animals, with purpose of diagnosis, treatment, mitigation, or prevention of any disease or disorder. Parenteral administration is excluded^15^.

B Another commonly utilized assay is the bioactive compound(s) disk diffusion assay, which is a useful screening tool, but has the drawback of the unknown diffusion factor for NPs, which can become a limiting factor when searching for synergistic effects, *e.g*., yielding false negative results. In some cases, a pre-screening panel was performed applying the agar diffusion test. It should be noted that deviations or disagreement between the results of those two methods can be observed, in some cases due to the low solubility or low diffusion of the analyte^22^.

C The term **natural products *sensu lato***, as applied in here, includes extracts derived from living organisms as well as purified compounds from such an extracts. These compounds could be obtained from either the direct original source, as biotechnological products, or from organic synthesis, being commercially available, or isolated at laboratory scale.

D For instance, the associated toxicities to the most frequently studied antibiotics are: *i)* ampicillin can induce the development of asthma^28,29^; *ii)* ciprofloxacin can affect the circulatory system in 1% of the cases, and tendinitis^29^. Also, occasional anaphylactic reactions due to beta–lactamics administration have been reported^28,30^. *iii)* aminoglycosides (*e.g*., gentamicin) can cause ototoxicity [5% –10%], which can affect, adults, children and fetus; they can interact with other medications enhancing the toxicity effects^28,31^.

E Non–polar extracts were usually redissolved in dimethylsulfoxide (DMSO), and then tested for their bioactivity. DMSO is not present in the final drug formulations, but it is commonly utilized to dissolve low polarity compounds in aqueous systems, instead of non-polar solvents such as hexane. DMSO by itself has a metabolic effect in cells that can affect the outcome bioactivity results^44^. Control experiments quantifying solvents effects were not generally clearly stated in the studies summarized in Tables 1 and 2. This type of experiments could even be considered as a ternary mixture: NPs, CAA, and the solvent. Other approaches for suspension or dissolution of NPs are needed in order to reduce false positives or potency overestimation of a given NPs.

## Notes

### Competing Interest Statement

The authors have declared no competing interest.

### Funding Statement

No external funding was received for this study.

### Author Declarations

The project did not involve human subjects.

